# Spatial Inequities in COVID-19 Testing, Positivity, Confirmed Cases and Mortality in 3 US Cities: an Ecological Study

**DOI:** 10.1101/2020.05.01.20087833

**Authors:** Usama Bilal, Loni P. Tabb, Sharrelle Barber, Ana V. Diez Roux

## Abstract

**Background:** Preliminary evidence has shown inequities in COVID-19 related cases and deaths in the US.

**Objective:** We explored the emergence of spatial inequities in COVID-19 testing, positivity, confirmed cases, and mortality in New York City, Philadelphia, and Chicago during the first six months of the pandemic.

**Design:** Ecological, observational study at the zip code tabulation area (ZCTA) level from March to September 2020.

**Setting:** Chicago, New York City and Philadelphia.

**Participants:** All populated ZCTAs in the three cities.

**Measures:** Outcomes were ZCTA-level COVID-19 testing, positivity, confirmed cases, and mortality cumulatively through the end of September. Predictors were the CDC social vulnerability index and its four domains, obtained from the 2014-2018 American Community Survey. We examined the spatial autocorrelation of COVID-19 outcomes using global and local Moran’s I and estimated associations using spatial conditional autoregressive negative binomial models.

**Results:** We found spatial clusters of high and low positivity, confirmed cases and mortality, co-located with clusters of low and high social vulnerability. We also found evidence for the existence of spatial inequities in testing, positivity, confirmed cases and mortality for the three cities. Specifically, neighborhoods with higher social vulnerability had lower testing rates, higher positivity ratios, confirmed case rates and mortality rates.

**Limitations:** ZCTAs are imperfect and heterogeneous geographical units of analysis. We rely on surveillance data, which may be incomplete.

**Conclusion:** We found spatial inequities in COVID-19 testing, positivity, confirmed cases, and mortality in three large cities of the US.

**Registration:** N/A

**Funding source:** NIH (DP5OD26429) and RWJF (77644)

## Introduction

As of the end of 2020, the COVID-19 pandemic had taken the lives of more than 1.5 people worldwide, while in the US deaths have surpassed 350,000 (1). Cities across the globe have emerged as especially vulnerable to COVID-19. Cities are characterized by diverse populations and are home to pronounced differences in health by race and socioeconomic position, often referred to as health inequities because they are avoidable and unjust(2). The presence of large racial and ethnic differences in COVID19 within US cities has already been documented. For example in New York City, both Blacks and Hispanics have double the age-adjusted mortality rate as compared to non-Hispanic whites(3), in Chicago 50% of deaths have occurred in Blacks, who make up only 30% of the population(4), while in Philadelphia, age-specific incidence, hospitalization, and mortality rates for Blacks and Hispanics are 2-3 times higher than for non-Hispanic whites(5). These stark differences by race are consistent with racial health inequities in many health outcomes and likely reflect multiple interrelated processes linked to structural inequity, historical racist policies, and residential segregation(6-8).

US cities are characterized by strong residential segregation by both race/ethnicity and income, one of the most visible manifestations of structural racism(9). Residential segregation results in stark differences across neighborhoods in multiple factors that could be related to both the incidence and severity of COVID-19, including factors related to transmission (e.g. overcrowding, jobs that do not allow social distancing) and factors related to severity of diseases (higher prevalence of chronic health conditions related to neighborhood environments, greater air pollution exposures and limited access to quality health care)(6-8,10,11). Few studies have systematically characterized spatial inequities in COVID related outcomes in cities over the course of the pandemic.

Characterizing social and spatial inequities in cities is critical to developing appropriate interventions and policies to prevent COVID-19 deaths in the future and mitigate economic and racial inequities. We used data from three large US cities, Chicago, New York City, and Philadelphia, to characterize spatial and social inequities in testing, positivity, confirmed cases, and mortality.

## Methods

### Setting

We used data on total numbers of tests, confirmed cases, and deaths by zip code tabulation area (ZCTA) of residence from Chicago, New York City (NYC), and Philadelphia. For Chicago, we downloaded data from the Chicago Department of Public Health(12), including cumulative data from through October 3^rd^, 2020. For NYC, we downloaded cumulative data made available by NYC Department of Health and Mental Hygiene in their GitHub repository(13) through October 1^st^, 2020. For Philadelphia, we downloaded data from the Philadelphia Department of Public Health made available in OpenDataPhilly (5), including cumulative data through October 1^st^ (5).

### Outcomes

Study outcomes included four COVID-19 indicators: (1) testing rates (total tests/population); (2) positivity ratio (14) (confirmed cases/total tests); (3) confirmed case rates (confirmed cases/population); and (4) mortality rates (deaths/population). For all indicators, we computed rates cumulatively through the end of the period.

### Predictors

To obtain a summary of social conditions in each area of residence we used the 2018 CDC’s Social Vulnerability Index (SVI)(15). Recent research has found the SVI is predictive of COVID-19 incidence and mortality at the county level(16). The SVI reflects the community’s ability to prevent human suffering and financial loss in the event of disaster, including disease outbreaks(15). It includes four domains: socioeconomic status, household composition & disability, minority status & language, and housing type & transportation, along with a summary score with all four domains. The four domains and summary score were calculated by the CDC at the census tract level using data from 15 variables from the 2014-2018 American Community Survey. Census tracts were ranked according to the values of each of the 15 variables, and percentile ranks were computed for each census tract within each state (in this case, Illinois, New York, and Pennsylvania). See **Appendix** for more details.

To aggregate the SVI to the ZCTA level, we used the Census Bureau’s ZCTA to Census Tract Relationship File, and computed a weighted mean of the SVI by ZCTA, using the population of the census tract in the ZCTA as the weight. A higher value of the SVI or of its component scores signifies higher vulnerability, either overall or in its four domains. For example, a higher vulnerability in the “socioeconomic status” domain reflects a higher proportion of people living in poverty, unemployed, with lower income or without a high school diploma. A higher vulnerability in the “housing type & transportation” domain reflects a higher number of people living in multi-unit structures, mobile homes, in crowded situations, without a vehicle, or living in group quarters.

Since the SVI represents the rank of each census tract (or ZCTA) within each state, we transformed the SVI to make coefficients comparable across cities. We first excluded all ZCTAs that were not part of each city, and then standardized SVI and its domain scores by subtracting the mean and dividing by the standard deviation (SD) for each city separately.

### Analysis

We conducted our analysis in three steps. First, we explored the spatial distribution of each of the five predictors (four domains and summary score) and the four outcomes (testing, positivity, confirmed cases and mortality) cumulative through the end of September, using choropleth maps. To explore whether there was spatial autocorrelation, we computed global Moran’s I(17,18). To show the location of spatial clusters, we computed the local indicator of spatial association (LISA) or local Moran’s I(17,18) and display clusters with a p-value<0.05.

Global Moran’s I statistic estimates the overall degree of spatial correlation, i.e., the degree to which the ZTCA rates of interest, such as testing, tend to be geographically located close together, far apart, or distributed randomly across the larger area, in this case each city. Moran’s I ranges from −⍰ 1 to 1, with positive values suggesting spatial autocorrelation and negative values suggesting dispersion (i.e., similar rates located far from each other), and values close to 0 indicating a random distribution. Significant p-values indicate evidence of departures from complete randomness.

The local Moran’s I index for each individual ZTCA reflects similarity of rates with those of nearby areas and can help identify outliers. These local indexes are relative measures with dimensions interpretable only by z-scores and their associated p-values. Hot (cold) spots are contiguous areas with consistently high (low) testing rates, while outliers would, for example, be areas with high (or low) testing rates surrounded primarily by areas with lower (or higher) testing rates. Here, we refer to hot and cold spots as high-high and low-low areas, respectively, and to outliers as high-low and low-high areas.

Second, we examined the relations between SVI and each of the outcomes through the end of the study period using scatterplots and smoothed loess lines. Third, to estimate the strength of the association between each predictor and outcome we considered using a Poisson model. However, after exploring the distribution of the outcomes, and after checking for overdispersion in Poisson models using the approach by Gelman and Hill(19), we opted for a negative binomial model. Negative binomial models relax the assumption of equality between the mean and variance, allowing for overdispersion. We fitted a separate model for each city and included the five predictors (four domains and summary score) in separate models. To account for the role of age in determining testing practices, influencing the probability of transmission, and its causal role on mortality, we adjusted all models by the % of people aged 65 or above in the ZCTA.

To account for spatial autocorrelation of the outcomes, we fitted a Besag-York-Mollie (BYM)(20) conditional autoregressive model, including a structured and unstructured ZCTA random effect, both following an intrinsic Gaussian Markov random field (IGMRF)(20). The structured spatial random effect takes into consideration that ZCTAs are more similar to other neighboring ZCTAs as compared to those further away. We defined neighboring ZCTAs based on regions with contiguous boundaries, defined as sharing one or more boundary point. We fitted this model using integrated nested Laplace approximations (INLA), a method approximating Bayesian inference(20,21). While this approach is an approximation-based method, it has previously shown accuracy and minimizes computational time(20-22). Details on model specification are provided in the **Appendix**. Results are shown as rate ratios associated with a one SD higher value of the SVI or its domains, separately for each city.

All analyses were conducted using R v4.0.2(23). Spatial analysis were conducted using the R package spdep(24) and R-INLA(25). More details on data management, the social vulnerability index, and the models are available on **the Appendix**. Code for replication is available at: https://github.com/usamabilal/COVID_Disparities

This study was funded with support from the NIH under grant DP5OD26429 and the Robert Wood Johnson Foundation under grant RWJF 77644. The funders had no role in study design, data collection and analysis, decision to publish, or preparation of the manuscript

## Results

We included a total of 58, 177, and 46 zip codes in Chicago, NYC and Philadelphia, respectively. From the beginning of the outbreak up to the latest available date (October 3^rd^ in Chicago and October 1^st^ in NYC and Philadelphia), a total of 674,929, 2,383,919, and 411,559 tests had been conducted in Chicago, New York City and Philadelphia, respectively. There were 81,657, 233,397, and 37,307 confirmed cases, and 2974, 19,149, and 1,803 COVID-19 deaths, respectively.

We found that testing, positivity, confirmed cases, and mortality were spatially autocorrelated in the three cities (global Moran’s I ranging from 0.198 to 0.803, p<0.001 in all cases except confirmed cases in Philadelphia, with p=0.011, for the null hypothesis of no spatial autocorrelation), with the exception of mortality in Philadelphia for which we did not find evidence for significant spatial autocorrelation (global Moran’s I=0.062, p=0.140). These patterns held after taking into consideration the spatial distribution of the SVI (global Moran’s I ranging from 0.127 to 0.705, p<0.05 in all cases), with the exception of confirmed cases and mortality for Philadelphia, that did not show significant spatial autocorrelation after controlling for the SVI (Moran’s I=-0.011 and −0.058, p=0.440 and 0.630). The **Appendix** shows global Moran’s I and associated Moran’s scatterplots for the three cities and four outcomes.

**Figures 1-3** shows the spatial patterning of clusters of testing, positivity, confirmed cases, mortality, and social vulnerability in Chicago, New York City, and Philadelphia, respectively. Generally, clusters of high positivity and confirmed cases were spatially co-located with clusters of high social vulnerability.

**Figure 1:**
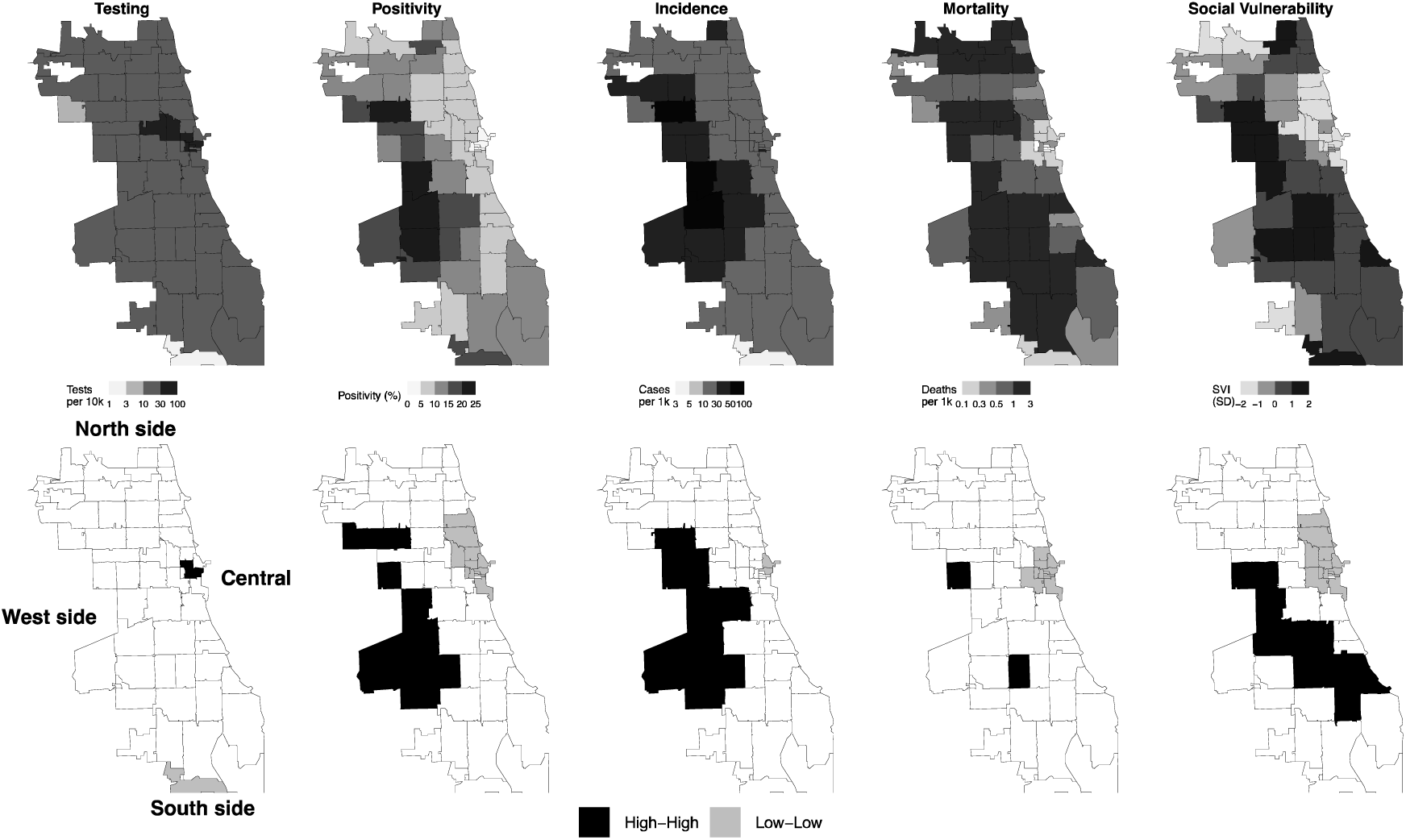
Spatial distribution and clusters of COVID-19 testing, positivity, confirmed cases, mortality, and social vulnerability in ZCTAs of Chicago. <u>Footnote</u>: clusters calculated using local Moran’s I statistic; clusters have a p-value <0.05. SD: standard deviation.

**Figure 2:**
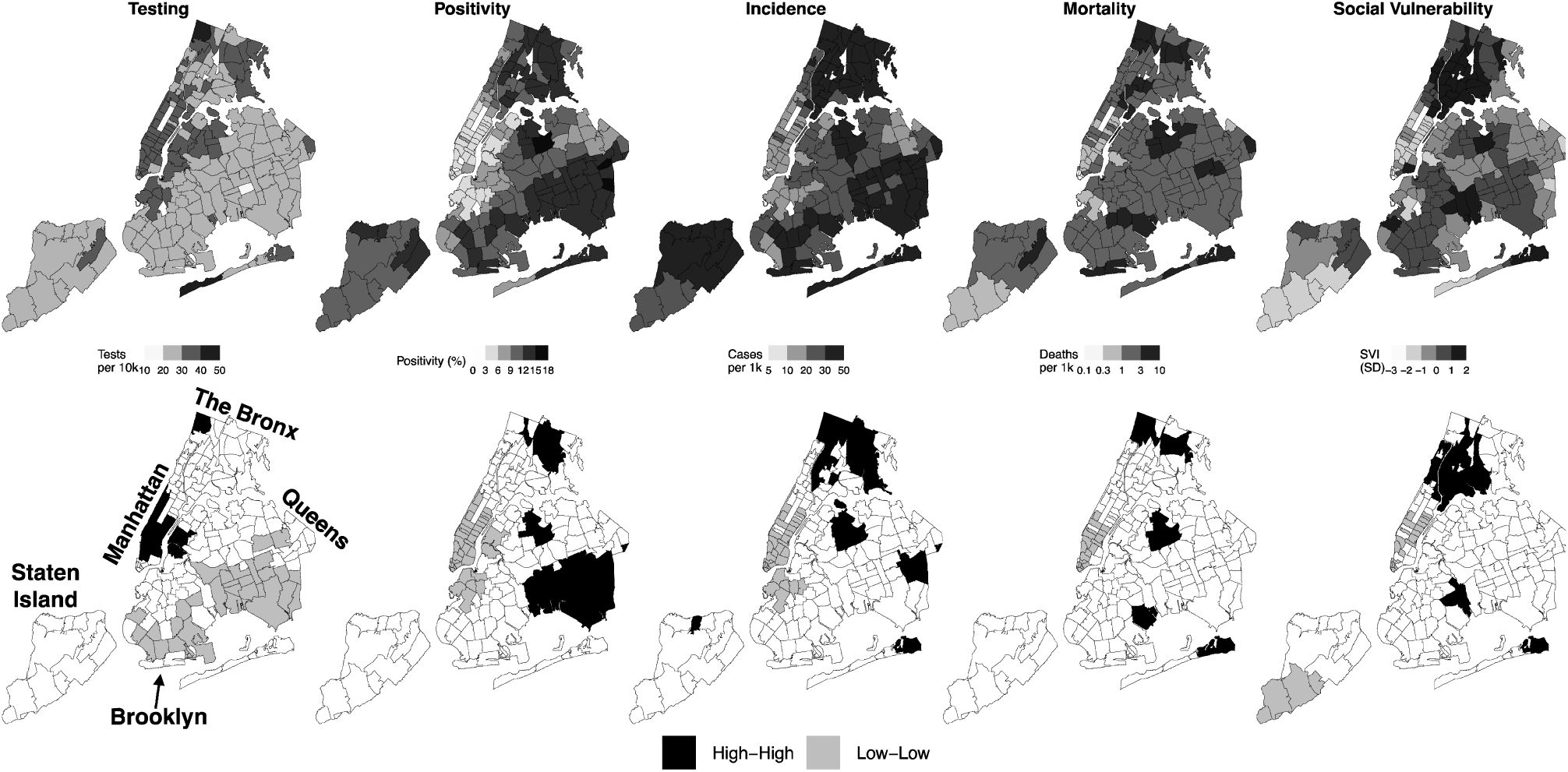
Spatial distribution and clusters of COVID-19 testing, positivity, confirmed cases, mortality, and social vulnerability in ZCTAs of New York City. <u>Footnote</u>: clusters calculated using local Moran’s I statistic; clusters have a p-value <0.05. SD: standard deviation.

**Figure 3:**
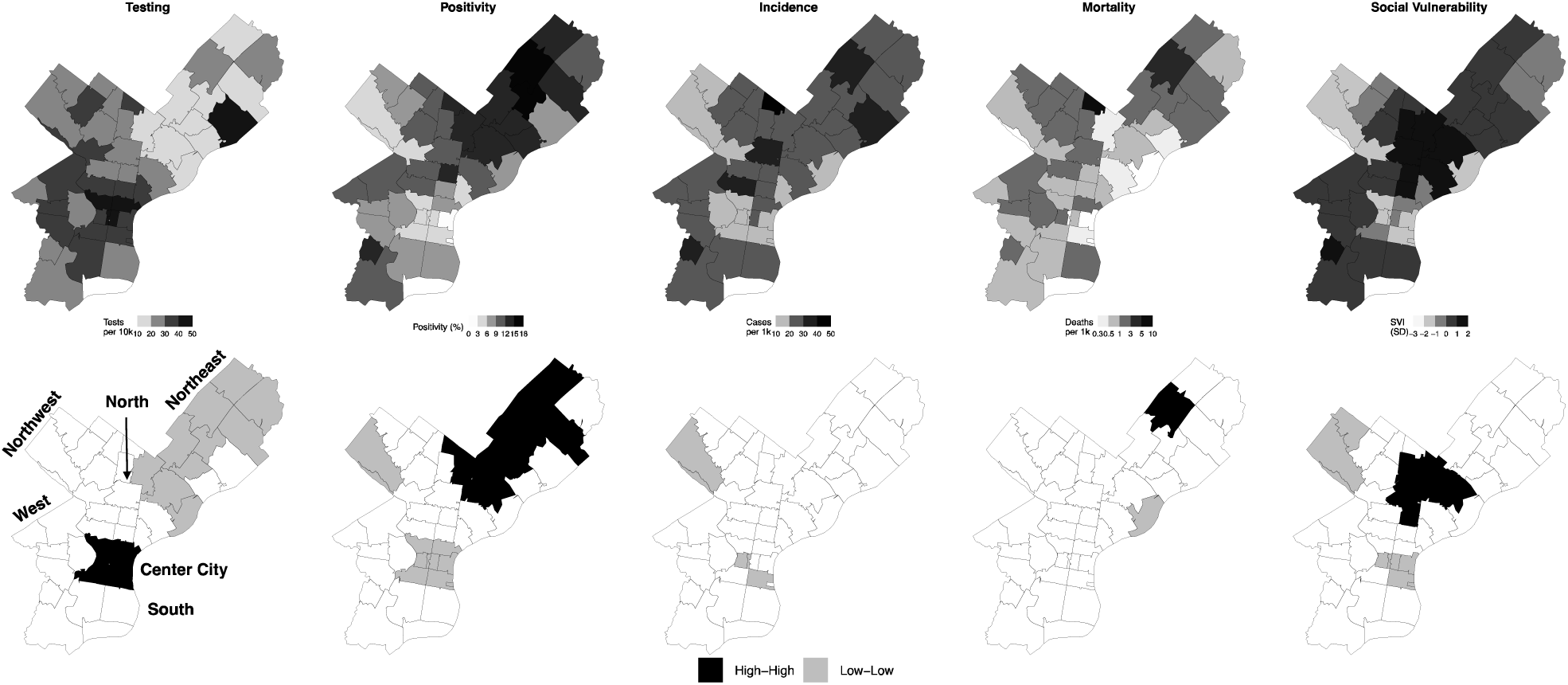
Spatial distribution and clusters of COVID-19 testing, positivity, confirmed cases, mortality, and social vulnerability in ZCTAs of Philadelphia. <u>Footnote</u>: clusters calculated using local Moran’s I statistic; clusters have a p-value <0.05. SD: standard deviation.

Areas of the West and South sides of Chicago have clusters of high positivity, confirmed cases and mortality (**Figure 1**). For example, ZCTAs 60636 and 60644 in the South and West Side, respectively, are significant clusters of high positivity (local Moran’s I=5.32 and 2.81, p<0.001 and =0.018), confirmed cases (local Moran’s I=5.55 and 3.65, p=0.007 and 0.013), and mortality (local Moran’s I=4.61 and 3.26, p=0.020 and 0.024). Conversely, the Central and North sides of Chicago had clusters of low positivity, confirmed cases and mortality, along with high testing (**Figure 1**). For example, ZCTA 60601 in Central Chicago is a cluster of high testing, low positivity, confirmed cases and mortality (local Moran’s I=5.50, 9.53, 5.88 and 6.78, p=0.003, 0.004, <0.001 and 0.001), while ZCTA 60661 in the North side is a cluster of low positivity and mortality (local Moran’s I=4.09 and 6.22, p=0.015 and <0.001).

In NYC, there were clusters of high positivity, confirmed cases and mortality in The Bronx and Queens (**Figure 2**). For example, ZCTAs 10467 and 11368 in The Bronx and Queens are statistically significant clusters of high positivity (local Moran’s I=6.02 and 6.36, p=0.019 and 0.010), confirmed cases (local Moran’s I=14.2 and 11.46, p<0.001) and mortality (local Moran’s I=6.17 and 13.54, p=0.016 and <0.001). There were also a number of clusters of high testing and low positivity, confirmed cases and mortality in Manhattan and the adjacent areas of Brooklyn (**Figure 2**). For example, ZCTA 10014 in Manhattan is a cluster of high testing, low positivity, confirmed cases and mortality (local Moran’s I=7.08, 7.15, 8.71 and 4.10, p<0.001, <0.001, <0.001 and =0.008), while ZCTA 11238 in Brooklyn is a cluster of low positivity and confirmed cases (local Moran’s I=6.97 and 5.17, p=0.009 and <0.001).

In Philadelphia, we found clusters of high testing and low positivity and confirmed cases in Center City, including ZCTAs 19102 (local Moran’s I=18.87, 3.90 and 11.04, p<0.001, <0.001 and =0.037) and 19103 (local Moran’s I=5.44, 5.35, and 7.341, p=0.002, 0.002 and <0.001). Most of North and Northeast Philadelphia was contained in a cluster of low testing and high positivity. For example, ZCTAs 19124 and 19149 are significant clusters of low testing (local Moran’s I=8.02 and 4.43, p<0.001 and =0.009) and high positivity (local Moran’s I=5.51 and 7.70, p=0.010 and <0.001).

We visually explored the relationship between the SVI and cumulative testing rates, positivity ratios, confirmed case rates and mortality rates for the three cities (**Figure 4**). Testing rates were slightly lower in areas of higher vulnerability in Chicago, NYC and Philadelphia. Positivity, confirmed cases, and mortality all increased monotonically with increasing social vulnerability in Chicago. A similar pattern was observed in NYC, but the increase was less marked in ZCTAs above mean vulnerability. Similar patterns were observed in Philadelphia for positivity and confirmed cases but the SVI was not consistently associated with mortality.

**Figure 4:**
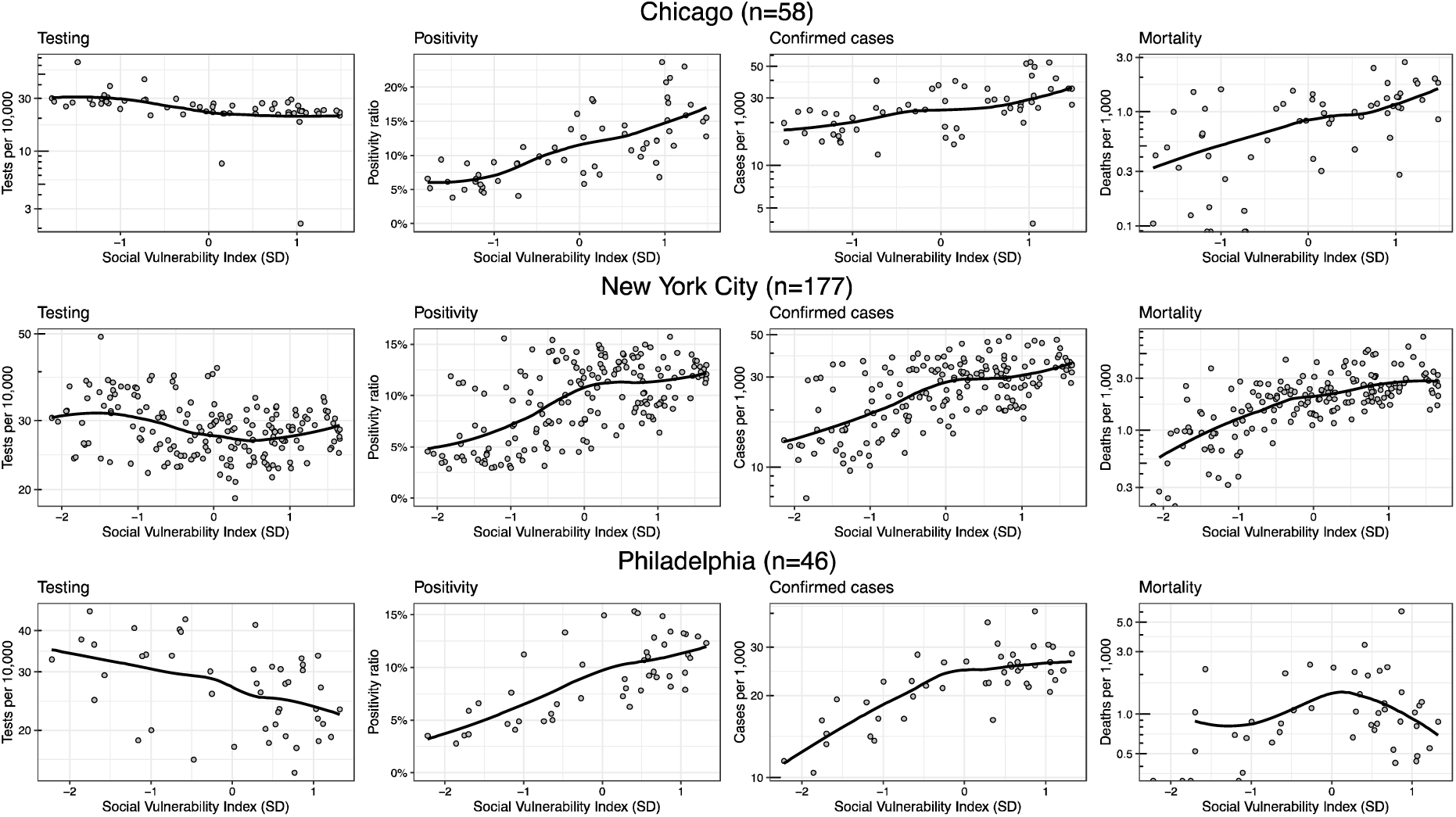
Scatterplots showing the relationship between the social vulnerability index and COVID-19 testing, positivity, confirmed cases and mortality at the ZCTA-level in Chicago, NYC, and Philadelphia. <u>Footnote</u>: solid lines are loess smoothers for each city separately. **The SVI has been standardized for each city, so that its units are standard deviations (SD)** of the SVI, for each city separately.

**Table 1** shows rate ratios of each outcome (cumulatively across the full study period), associated with a one SD higher value of the SVI index and its four domains, after adjusting for the % of the population aged 65 or above. A higher social vulnerability was associated with 13%, 3%, and 9% lower testing rates in the three cities, although confidence intervals crossed the null. Associations of SVI with positivity, confirmed cases, and mortality were similar in the three cities. A 1 SD higher SVI was associated with 40%, 37% and 40% higher positivity in Chicago, NYC and Philadelphia, 22%, 33%, and 27% higher confirmed cases, and 44%, 56%, and 58% higher mortality. For the three cities, we found that the social vulnerability domains of socioeconomic status, household composition & disability, minority status & language were associated with the study outcomes similarly to the overall index. However, weaker or even opposite associations were observed for the housing type & transportation component.

**Table 1:** Relative rates of testing, positivity, confirmed cases, and mortality. associated with zip code tabulation area (ZCTA) social vulnerability index (SVI) and its four domains in Chicago, NYC, and Philadelphia.

| City | Variable | Testing | Positivity | Incidence | Mortality |
| --- | --- | --- | --- | --- | --- |
| Chicago | **Social Vulnerability Index** | **0.87(0.74;1.01)** | **1.40(1.25;1.58)** | **1.22(1.04;1.42)** | **1.44(1.15;1.80)** |
|  | Socioeconomic Status | 0.87(0.74;1.02) | 1.46(1.30;1.65) | 1.27(1.09;1.49) | 1.52(1.21;1.91) |
|  | Household composition & disability | 0.86(0.74;1.02) | 1.34(1.16;1.55) | 1.17(0.98;1.39) | 1.46(1.14;1.87) |
|  | Minority status & language | 0.95(0.82;1.09) | 1.34(1.20;1.49) | 1.27(1.11;1.45) | 1.33(1.08;1.64) |
|  | Housing type & transportation | 0.95(0.82;1.11) | 0.95(0.82;1.10) | 0.89(0.76;1.05) | 0.91(0.71;1.16) |
| New York City | **Social Vulnerability Index** | **0.97(0.94;1.00)** | **1.37(1.29;1.46)** | **1.33(1.26;1.41)** | **1.56(1.46;1.67)** |
|  | Socioeconomic Status | 0.94(0.91;0.97) | 1.47(1.39;1.56) | 1.39(1.32;1.46) | 1.62(1.51;1.73) |
|  | Household composition & disability | 0.98(0.96;1.01) | 1.35(1.27;1.42) | 1.32(1.26;1.38) | 1.39(1.30;1.48) |
|  | Minority status & language | 0.94(0.91;0.97) | 1.36(1.28;1.45) | 1.28(1.21;1.35) | 1.51(1.41;1.63) |
|  | Housing type & transportation | 1.10(1.07;1.12) | 0.84(0.78;0.90) | 0.92(0.86;0.98) | 1.01(0.93;1.10) |
| Philadelphia | **Social Vulnerability Index** | **0.91(0.82;1.02)** | **1.40(1.25;1.55)** | **1.27(1.15;1.42)** | **1.58(1.24;2.00)** |
|  | Socioeconomic Status | 0.90(0.80;1.01) | 1.41(1.26;1.59) | 1.27(1.14;1.43) | 1.49(1.16;1.91) |
|  | Household composition & disability | 0.90(0.80;1.00) | 1.37(1.23;1.53) | 1.23(1.10;1.37) | 1.31(1.03;1.67) |
|  | Minority status & language | 0.94(0.85;1.05) | 1.26(1.13;1.42) | 1.20(1.08;1.33) | 1.47(1.13;1.91) |
|  | Housing type & transportation | 1.03(0.93;1.15) | 1.01(0.88;1.15) | 1.03(0.92;1.16) | 1.28(1.02;1.60) |

**Footnote:** Rate ratios (95% CrI) for a 1 SD higher value of the SVI and its four domains. All models are adjusted for the % of the population in the ZCTA aged 65 or above. Data is cumulative through October 1<sup>st</sup>, 2020 (NYC and Philadelphia) and October 3<sup>rd</sup>, 2020 (Chicago).

## Discussion

We documented large spatial inequities in COVID-19 through the end of September 2020 in three large US cities, Chicago, NYC and Philadelphia, with more vulnerable neighborhoods having a higher positivity, confirmed cases and mortality, and lower testing rates. We also found clusters of high and low positivity, confirmed cases, and mortality, co-located with areas of high and low vulnerability, respectively. Notably we observed very strong inequities in mortality, with mortality rates increasing by ∼50% for each SD higher SVI index.

Findings from this study are consistent with other studies that have examined inequities in COVID-19 incidence by ZCTA in other cities. For example, Chen & Krieger reported a monotonic increase in confirmed cases in ZCTAs of Illinois and NYC with decreasing levels of area-level socioeconomic status(10). Analysis at the county level by the same authors showed similar gradients(10), consistent with other research(26,27), including a study using the SVI as a predictor at the county level(16). We found that, within these large cities, clusters of high and low positivity and confirmed cases that were mostly co-located with clusters of high and low vulnerability, respectively. These include areas of concentrated poverty and with a history of extreme racial segregation(7), including West and North Philadelphia, the West Side of Chicago, and The Bronx in NYC. Notably Chicago, Philadelphia, and NYC are among the top 10 most segregated cities in the country (28).

As others have noted(6-8) potential explanations for neighborhood inequities in incidence may include differential exposure to the virus and as well as differential susceptibility to infection. Residents of higher SVI neighborhoods likely have higher exposure to the virus because of the types of jobs they have (such as essential workers within the healthcare, personal care, production, or service industries(29), personal care or service occupations(30)), lack of telecommuting options(31), dependence on mass transit use(32), and because of overcrowding within households(33). Whether there are factors associated with differential susceptibility to infection is still unclear, but prior research on respiratory viruses has documented that stress linked to disadvantage may increase the likelihood of developing disease after exposure(34,35).

We also found narrow inequities in testing, accompanied by wider inequities in positivity. The inequities in positivity suggest that, despite apparently small inequities in testing, testing may actually be lower than it needs to be in neighborhoods with high SVI and possibly higher incidence. Barriers to testing can include unequal location of testing sites(36), lack of vehicle ownership(37), lack of health insurance(38), a usual source of care for referrals(39), and potential mistrust of the medical system(40). It is possible that the social patterning of infection has been changing over time as the pandemic progressed, beginning in wealthier areas (possibly linked to business travel(41)) and subsequently shifting to more deprived areas. As more longitudinal data becomes available, understanding longitudinal patterns may help in the preparedness for future outbreaks. Our ability to adequately characterize inequities in incidence necessarily requires equal access to testing.

A major finding was the substantially higher mortality rate in neighborhoods with a higher SVI. Vulnerability to severe disease and death by COVID-19 are related to the presence of previous comorbidities, such as cardiovascular disease, diabetes and hypertension(42). Since these comorbidities are more prevalent in people of lower socioeconomic status and racial/ethnic minorities(43,44), it is expected that, at equal levels of exposure, these groups will suffer more severe consequences from COVID-19. Other factors may also affect the severity of disease and the case-fatality rates including access to and quality of health care and the role of other factors including co-occurring social factors (e.g. stressors) and environmental factors (e.g. air pollution). In fact, a study with 17 million records in the UK has shown that, even after adjusting for a number of comorbidities, racial/ethnic minorities and people living in socioeconomically deprived areas had a higher risk of death after infection(42). However, two recent studies using data from Michigan and the Veteran Affairs health system suggest that inequities in mortality are driven by differences in infection rates, rather than differential vulnerability(45,46). In our study, we found that the relative risks of mortality associated with higher zip code social vulnerability were slightly higher than those observed for confirmed cases, but underestimation of the underlying incidence in higher SVI neighborhoods (because of lower testing) could partly explain this difference.

We found that the domain of social vulnerability due to housing type & transportation showed inconsistent associations as compared to the other domains or the overall summary social vulnerability index. This domain includes variables detailing the proportion of the population living in multi-unit structures, mobile homes, group quarters, in crowded situations, or without a vehicle. It is possible that these variables do not relate to differences in COVID-19 outcomes within cities because either they are not heterogeneous enough or they simply do not capture true underlying determinants.

An important limitation of our study is the likely underestimation of inequities in confirmed case rates due to the lack of systematic widespread testing. We also lack individual-level data, and rely on aggregated surveillance data. In addition, ZCTAs are very imperfect proxies for neighborhoods(47). Heterogeneity in the sociodemographic composition within zip codes may have led to underestimation of inequities(48). However, zip codes represent easy-to-collect data in the middle of a public health emergency when more detailed geocoding is less available.

## Conclusion

We found large spatial inequities in COVID-19 testing, positivity, confirmed cases and mortality in three large cities of the US and strong associations of COVID-19 positivity, confirmed cases, and mortality with higher neighborhood social vulnerability. These within-city neighborhood differences in COVID-19 outcomes emerge from differences across neighborhoods generated and reinforced by residential segregation linked to income inequality and structural racism(49-51), coupled with decades of systematic disinvestment in segregated neighborhoods(7-10,50,52). Addressing these structural factors linked to income inequality, racism and segregation will be fundamental to minimizing the toll of the pandemic but also to promoting population health and health equity across many other health conditions.

## Data Availability

Data is publicly available. Code is available on a github repository.

https://github.com/usamabilal/COVID_Disparities

## Reproducible research statement

<u>Protocol</u>: not available

<u>Statistical Code</u>: Available at https://github.com/usamabilal/COVID_Disparities

<u>Data</u>: Available at https://github.com/usamabilal/COVID_Disparities

## Acknowledgements

The authors want to acknowledge help by Alyssa Furukawa on data collection, and Dr. Rene Najera for useful code to calculate spatial autocorrelation.

## Preprint server

This article was published and updated on medRxiv on February 1st, 2021: https://doi.org/10.1101/2020.05.01.20087833

