## Appendix for "Spatial Inequities in COVID-19 Testing, Positivity, Confirmed Cases and Mortality in 3 US Cities: an Ecological Study"

**Appendix index:**

- **Appendix 1: data sources**
- **Appendix 2: social vulnerability index and its components**
- **Appendix 3: exploration for appropriateness of Poisson model**
- **Appendix 4: exploration of spatial autocorrelation and appropriateness of non-spatial models**
- **Appendix 5: local spatial autocorrelation and cluster detection**
- **Appendix 6: INLA model specification**

**Appendix 1: Data sources and management**

- **Chicago:** we downloaded the following spreadsheet on 10/07
  - **URL:** <https://data.cityofchicago.org/Health-Human-Services/COVID-19-Cases-Tests-and-Deaths-by-ZIP-Code/yhhz-zm2v>
- **New York City:** we downloaded the following file corresponding to the October 1^st^ snapshot
  - **URL:** <https://github.com/nychealth/coronavirus-data/blob/697dcb6c9bd3531b3838a733bfb26aeb97538224/data-by-modzcta.csv>
- **Philadelphia:** we downloaded the counts of tests, cases and deaths on October 1^st^ from this link (note: cumulative historical snapshots are also available in the link below)
  - **URL:** <https://www.opendataphilly.org/dataset?q=covid&sort=metadata_modified+desc>
- **Social Vulnerability Index:** we downloaded the SVI and its components for Illinois, New York and Pennsylvania from the CDC’s Agency for Toxic Substances and Disease Registry website.
  - **URL:** [**https://svi.cdc.gov/data-and-tools-download.html**](https://svi.cdc.gov/data-and-tools-download.html)
- **Shapefiles:** we downloaded shapefiles for ZCTAs and other required levels from the census cartography files website.
  - **URL:** [**https://www.census.gov/geographies/mapping-files/time-series/geo/carto-boundary-file.html**](https://www.census.gov/geographies/mapping-files/time-series/geo/carto-boundary-file.html)
  - **Note:** for NYC we used the shape files provided by NYC DOH [**https://github.com/nychealth/coronavirus-data/tree/master/Geography-resources**](https://github.com/nychealth/coronavirus-data/tree/master/Geography-resources)
- **Other census variables (e.g. total population for the ZCTA):** we downloaded data from the 2014-2018 ACS using the R package tidycensus (see code).
  - **Note:** for NYC we used the improved population estimates provided by NYC DOH in their COVID-19 files (see above).
- **The resulting final files are available with the rest of the code at** [**https://github.com/usamabilal/covid_disparities**](https://github.com/usamabilal/covid_disparities)

**Appendix 2: social vulnerability index and its components**

The social vulnerability index is a measure of social vulnerability, defined as “The degree to which a community exhibits certain social conditions, including high poverty, low percentage of vehicle access, or crowded households, may affect that community’s ability to prevent human suffering and financial loss in the event of disaster.”(1).

The index was created by summing the percentiles of the distribution of a number of variables in four domains. The overall ranking was created by adding up the rankings of the four domains. A higher value of the index reflects higher social vulnerability, either overall or in each of the four domains.

| **Domain** | **Variable** |
| --- | --- |
| **Socioeconomic Status** | Below Poverty |
|  | Unemployed |
|  | Income |
|  | No High School Diploma |
| **Household Composition & Disability** | Aged 65 or Older |
|  | Aged 17 or Younger |
|  | Civilian with a Disability |
|  | Single-Parent Households |
| **Minority Status & Language** | Minority |
|  | Speaks English “Less than Well” |
| **Housing Type & Transportation** | Multi-Unit Structures |
|  | Mobile Homes |
|  | Crowding |
|  | No Vehicle |
|  | Group Quarters |

All data was obtained from the 2014-2018 5-year American Community Survey (ACS).

We used the state-level datasets for New York, Illinois and Pennsylvania, as recommended by CDC for all analysis that do not intend to make national comparisons.

**Appendix 3: exploration for appropriateness of Poisson model**

As indicated in the main manuscript, and to check for the appropriateness of a Poisson model to model our outcomes, we explored the mean and variance of the number of tests, confirmed cases and deaths in each city. We also fitted a Poisson model (non-spatial) including social vulnerability (or its domains) as a covariate, and checked for overdispersion, using the test suggested in Gelman & Hill(2).

Overall, we found that all outcomes had a much higher variance as compared to their means in every city, and we were also able to reject the null hypothesis of no overdispersion for each city/exposure/outcome combination (p<0.001 in all cases). For this reason, we opted to use negative binomial models to relax the assumption of no overdispersion.

**Appendix 4: exploration of spatial autocorrelation and appropriateness of non-spatial models**

As indicated in the main manuscript, we checked for the presence of global spatial autocorrelation using global Moran’s I(3-5). First, we ran Global Moran’s I on the four outcomes (testing rates, positivity, confirmed case rates, and mortality rates). Second, we ran global Moran’s I on the Pearson residuals of a (non-spatial) negative binomial model adjusted for the SVI. Third, we created a correlogram on the residuals of this negative binomial model.

Global Moran’s I compares the values of each ZCTA to the spatially weighted values of its neighboring ZCTAs (also called spatially lagged values)(5), aggregates these comparisons producing the global Moran’s I statistic that is then compared to the observed statistic under the null hypothesis of no spatial autocorrelation(5).	For all global Moran’s I tests, we used the moran.mc function from the spdep package. This function uses a permutation bootstrap test, in which values are assigned at random to ZCTAs, and the test statistic is computed for each permutation(5). We run this test using 9999 permutations.

We found that we could reject the null hypothesis of no spatial autocorrelation for all outcome/city combinations, with the exception of mortality in Philadelphia. These patterns persisted after controlling for the SVI, with one exception (confirmed cases in Philadelphia showed no spatial autocorrelation after controlling for the SVI)

**Global Moran’s I (p-value) for each outcomes by city**

| **Model** | **Variable** | **Chicago** | **New York City** | **Philadelphia** |
| --- | --- | --- | --- | --- |
| **Unadjusted** | Testing | 0.385 (<0.001) | 0.560 (<0.001) | 0.629 (<0.001) |
|  | Positivity | 0.625 (<0.001) | 0.803 (<0.001) | 0.674 (<0.001) |
|  | Confirmed cases | 0.431 (<0.001) | 0.691 (<0.001) | 0.198 (0.011) |
|  | Mortality | 0.370 (<0.001) | 0.380 (<0.001) | 0.062 (0.140) |
| **SVI Adjusted** | Testing | 0.127 (0.026) | 0.549 (<0.001) | 0.397 (<0.001) |
|  | Positivity | 0.439 (<0.001) | 0.705 (<0.001) | 0.486 (<0.001) |
|  | Confirmed cases | 0.244 (0.001) | 0.599 (<0.001) | -0.011 (0.430) |
|  | Mortality | 0.172 (0.017) | 0.253 (<0.001) | -0.058 (0.630) |

Footnote: Moran’s I calculated using the moran.mc function (9999 permutations); p-value in parenthesis for the null hypothesis of no spatial autocorrelation.

**Appendix 5: local spatial autocorrelation and cluster detection**

Global measures of autocorrelation, such as global Moran’s I above, are calculated from aggregations of relationships between local units, and can be broken down into their local components, producing local indicators of spatial association (LISA)(5). The local version of the Moran’s I takes each ZCTA-level comparison and tests them for divergence from the null hypothesis of no spatial autocorrelation(5).

Graphically, this is similar to plotting the values of each ZCTA (x-axis) vs the spatially lagged values of each ZCTA (y-axis); global Moran’s I statistic is then the slope of the linear fit of the spatially lagged values on the values of each ZCTA(5). Moreover, if we divide the Moran’s plot into four quadrants, the bottom-left and top-right quadrants represent observations with low values surrounded by areas of low values, and observations with high values surrounded by areas of high values, respectively (i.e., concordant relationships). The top-left and bottom-right quadrant represents observations of low values surrounded by areas of high values, and observations of high values surrounded by areas of low values, respectively (i.e., discordant relationships).

In the main manuscript, we map ZCTAs for which we reject the null of no spatial autocorrelation (p<0.05). Since we only found significant low-low and high-high areas (concordant relationships), we only map these two types of clusters in Figures 1-3. Below, we show a more detailed account of these values. Specifically, and for each city, we show Moran’s plot(5), and categorize each ZCTA according to its p-value.

**Moran’s plot for all four outcomes in the three cities of this study**

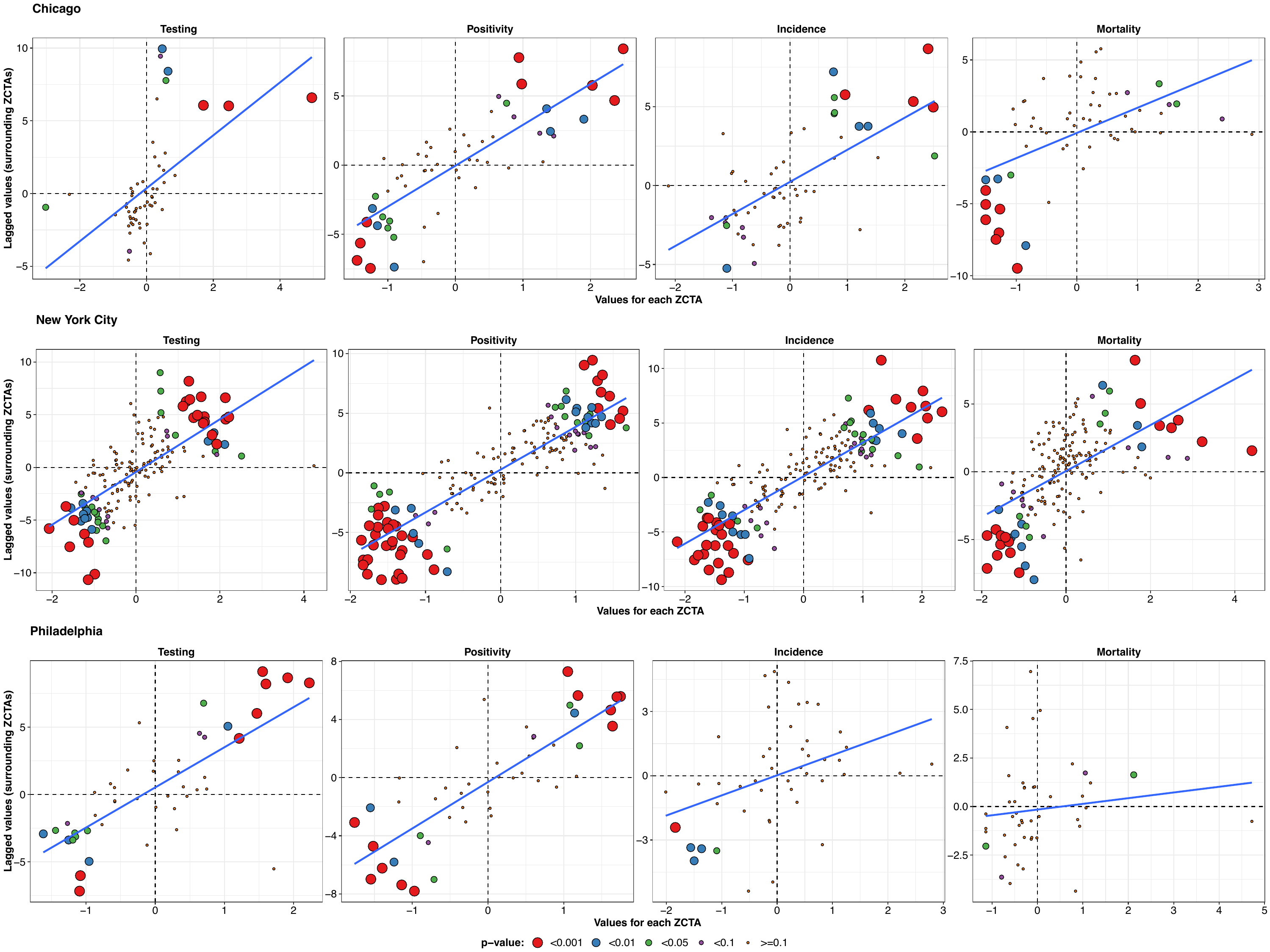

**Footnote**: p-values for each ZCTA correspond to the p-value of the local Moran’s I test of no divergence from the null of no spatial autocorrelation. Solid blue line corresponds to a linear fit of spatially lagged values ~ actual value of each ZCTA.

**Appendix 6: Model specification**

6.1: Model specification

We followed the model specification guidelines outlined in Blangiardo and Cameletti(6), specifically in sections 6.1.2 (BYM model) and 6.2 (Ecological regression). This model specification uses an intrinsic conditional autoregressive specification and an exchangeable random effect, resulting in the Besag-York-Molliè model(6), introducing two random effects (v and u): v is an area-specific random effect modelled as exchangeable, allowing for a correlation between units of the same city that is constrained to be the same across all units; u is the spatially structured random effect which takes into consideration that surrounding areas are more likely to be similar to each other than areas further away. As described in other sections of this Appendix, we opted to use a negative binomial model instead of a Poisson model, due to the overdispersed nature of our data.

6.2: Priors

All models assumed a rather non-informative prior for the hyperparameter precision and for the remaining parameters, rather than the default priors of the INLA R package(6-8). Specifically, we used a non-informative prior for the precision of the unstructured and spatial components of the BYM model, with Gamma (1, 0.5) as the distribution, as proposed in previous studies(9). For the coefficients of the fixed effects, we also assumed a non-informative prior of Normal(0, 10^3^), as proposed in previous studies(9). To check for the robustness of our findings to the choice of priors, we compared fixed effects and their 95% credible intervals from the model with the priors mentioned above compared to a model with the default priors of the INLA package. We found no major differences in fixed effects or their uncertainty estimates.

5.3: Convergence

Last, we assessed model convergence by exploring the Kullback-Leibler divergence (KLD) parameter for all coefficients of interest, finding it to be 0 or close to 0 in all cases, indicating good model convergence(10). Specifically, KLD < 10^^-5^ for all coefficients of the 60 spatial models (3 cities, 4 outcomes, 5 predictors).
